# A Bayesian Framework for Physiologically-Based Modeling of Flutter-Induced Aneurysm Progression

**DOI:** 10.64898/2026.02.09.26345810

**Authors:** Kiran Bhattacharyya

## Abstract

Current clinical risk stratification for thoracic aortic aneurysms (TAA) relies primarily on maximum diameter, which is a poor predictor of rupture. Recent fluid-structure interaction studies have identified a dimensionless “flutter instability parameter” (*N*_*ω*_ ) that accurately classifies abnormal aortic growth. However, this parameter currently serves as a static diagnostic snapshot. In this work, we propose a proof-of-concept computational framework that links flutter instability to microstructural tissue damage via a coupled system of ordinary differential equations (ODEs). We model a feedback loop where flutter-induced energy dissipation drives elastin degradation and collagen remodeling, which in turn reduces wall stiffness and amplifies the instability. To address the challenge of unobservable tissue properties, we implement a Bayesian inference engine to infer model parameters. We demonstrate feasibility on a synthetic patient cohort calibrated to published clinical growth rates and diameters. Our results show that this approach can infer hidden damage parameters and capture the qualitative bifurcation between stabilizing remodeling and runaway aneurysm expansion. While validation on real patient data remains essential, this work establishes the mathematical foundation for transforming a static physiomarker into a personalized prognostic trajectory.

## I. Introduction

Thoracic aortic aneurysm (TAA) is a silent but lethal condition characterized by the weakening and expansion of the aortic wall. Current guidelines recommend surgical intervention when the aortic diameter exceeds 5.5 cm or when the growth rate is ≥ 0.5 cm/year [1]. However, these geometric criteria are insufficient: many aneurysms rupture unexpectedly, while large ones remain stable for years. Moreover, measuring growth rates requires taking at least two measurements typically separated by 2–5 years, creating a lengthy diagnostic process with risk of aneurysm between measurements.

There is a critical need for biomechanical metrics that capture the underlying pathophysiology with measurements from a single point in time [2], [3]. Recently, Zhao et al. identified “blood-wall fluttering” as a potential driver of TAA progression [4]. They derived a dimensionless stability parameter, *N*_*ω*_, based on the Womersley number and wall stiffness which shows high diagnostic accuracy (AUC *>* 0.99). However, it does not model the *temporal* evolution of the disease.

Here, we bridge the gap between static identification and dynamic prognosis. We hypothesize a simple, mathematical model for a mechanistic feedback loop with a critical bifurcation [4], [5]: flutter instability transfers energy to the vessel wall, causing fatigue-induced degradation of the extracellular matrix (ECM). This degradation lowers wall stiffness, which in turn lowers the critical velocity for flutter, which can either 1) create a “vicious cycle” of accelerating growth or 2) lead to increasing collagen deposition that stiffens the aortic wall and stops further growth. Our approach integrates hyperelastic constitutive frameworks [6] with mechanobiological control theories [7], [8] with recent work that links hemodynamic instability to structural remodeling [4].

We present a proof-of-concept computational framework that:

1. Formalizes this mechanism into a coupled ODE system governing elastin and collagen dynamics.
2. Solves the inverse problem using Bayesian inference to estimate patient-specific damage rates from sparse clinical data.
3. Demonstrates, using synthetic patients calibrated to clinical statistics, the potential to predict “time-to-threshold” for surgical planning and the ability to categorize patients by risk.

## II. Methods

We propose a proof-of-concept computational framework that couples a model of arterial growth and remodeling with a fluid-structure interaction stability criterion. This forward model is embedded within a Bayesian inference framework to estimate patient-specific tissue degradation parameters from longitudinal clinical data. As real longitudinal datasets with paired hemodynamic and tissue mechanics measurements are scarce, we show feasibility using a synthetic patient cohort calibrated to published clinical statistics. We then use the fitted parameters to investigate the framework’s ability to capture the temporal evolution of representative disease states: stable, borderline, and unstable.

### A. Mathematical Framework

#### 1) The Flutter-Driven Damage Model

The core hypothesis of this work is that fluid-structure flutter instability drives aneurysm progression through fatigue-induced degradation of the aortic wall constituents. We model the evolution of the aortic wall state vector **y**(*t*) = [*D*(*t*), *ϕ*_*e*_(*t*), *ϕ*_*c*_(*t*)]^*T*^, where *D* is the vessel diameter, and *ϕ*_*e*_, *ϕ*_*c*_ are the volume fractions of elastin and collagen, respectively [9], [10].

The driving force for degradation is the *Damage Potential* Ψ(*t*), a dimensionless scalar quantifying the intensity of the flutter instability. We derive Ψ(*t*) from the “Flutter Instability Parameter” (*N*_*ω*_) proposed by Zhao et al. [4], which serves as a bifurcation parameter for the stability of pulsatile flow in a compliant tube:

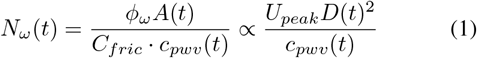

where *ϕ*_*ω*_ is the oscillatory flow acceleration, *A*(*t*) is the cross-sectional area, *c*_*pwv*_ is the pulse wave velocity (PWV), *C*_*fric*_ is a friction scaling factor derived from the Womersley number, and *U*_*peak*_ is the peak systolic blood velocity. Note that *U*_*peak*_ and *D* are directly measurable with MRI or ultrasound [4]. The damage potential is defined as a non-linear function of the excess instability above a critical threshold *N*_*thresh*_:

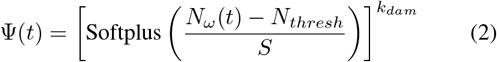

where *k*_*dam*_ is a damage exponent and *S* is a smoothing factor ensuring differentiability. We selected the Softplus function for its desirable mathematical properties: it provides a smooth, differentiable approximation to a threshold response (damage only accumulates when *N*_*ω*_ *> N*_*thresh*_), avoids numerical instabilities associated with hard thresholds, and introduces minimal additional parameters. While this formulation is phenomenological, it captures the essential concept that subthreshold flutter should not accumulate damage while superthreshold flutter drives degradation in a potentially nonlinear manner. The exponent *k*_*dam*_ allows the model to learn whether damage accumulation is linear (*k*_*dam*_ = 1) or accelerating (*k*_*dam*_ *>* 1) with respect to magnitude.

#### 2) Coupled Ordinary Differential Equations

The evolution of the state variables is governed by a system of coupled ODEs describing constituent turnover and geometric growth.

##### Elastin Degradation

Elastin is modeled as a non-regenerative structural protein in adults [9], [11]. Its depletion is driven by natural aging and flutter-induced fatigue:

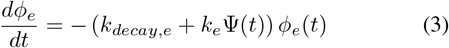

where *k*_*e*_ is the flutter-induced degradation rate constant estimated via inference and *k*_*decay,e*_ is the rate of elastin fraction reduction through aging inferred from published literature [10], [12]. The initial value of *ϕ*_*e*_(*t* = 0) is set to 0.3 for Bayesian estimation.

##### Collagen Remodeling

Collagen dynamics are modeled as a competition between stress-mediated synthesis and degradation. To prevent unbounded stiffening, we introduce a space-limitation term Γ(*ϕ*_*c*_) that damps synthesis as the collagen fraction approaches maximum and minimum values *ϕ*_*c,max*_ and *ϕ*_*c,min*_ (0.45, 0.05) [9]:

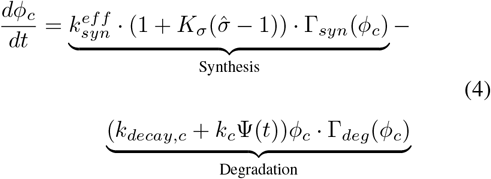

Here, 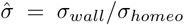 is the normalized wall stress ratio. The baseline synthesis rate 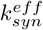 is calibrated to exactly balance natural decay *k*_*decay,c*_ at the homeostatic state 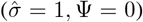. This formulation ensures that stable patients maintain wall integrity. The value for *k*_*decay,c*_ is derived from published literature [13], [14] while the values for *k*_*c*_ and *K*_*σ*_, a dimensionless collagen synthesis gain parameter [15], are estimated with inference. The initial value of *ϕ*_*c*_(*t* = 0) is set to 0.25 for Bayesian estimation.

##### Geometric Evolution

The diameter evolves according to a stress-growth law modulated by the effective wall stiffness *E*(*t*):

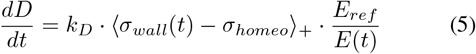

where *E*_*ref*_, the stiffness at *t* = 0, and *E*(*t*) are calculated via a rule-of-mixtures weighted by constituent fractions [7], [8], [16]. The stiffness values used for elastin and collagen were those reported in previous literature [17]–[19]. The feedback loop is closed as degradation reduces *E*(*t*), lowering 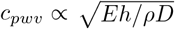 [20], which in turn increases *N*_*ω*_, creating a self-reinforcing cycle of instability. Note that *h* is the wall thickness which also reduces in proportion to the increase in diameter (initial value derived from prior work) [21].

### B. Synthetic Cohort Generation

Due to the scarcity of longitudinal aortic tissue mechanics data, we generated a synthetic patient cohort derived from the clinical growth rates reported in Zhao et al. [4].

#### 1) Data Extraction and Augmentation

Clinical data points comprising growth rates (cm/year) and binary stability classifications were digitized from the reference study (*N* = 48) [4]. To construct a full “Digital Twin” for each data point, we solved the inverse problem to assign physically consistent initial conditions:

- **Hemodynamic Assignment:** For patients in the unstable group (*N*_*ω*_ ≥ *N*_*ω,sp*_ and growth rate 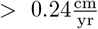 [4]), the peak initial systolic velocity 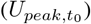 was assigned based on a noisy inversion of the growth law, ensuring that higher growth rates statistically correlated with higher peak systolic velocities preserving biological variance. We used a linear trend with Gaussian noise of *σ* = 0.15 m/s, approximately 10-20% of a reasonable baseline value.
- **Initial Geometry:** Baseline diameters 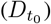 were sampled from age-dependent distributions for healthy and aneurysmal aortas [4]. Age was assigned randomly from a uniform distribution 𝒰 [30, 70].
- **Final Geometry:** We sampled the time in years for the second measurement after the initial measurement (*t*_*F*_ ) from a normal distribution with mean and standard deviation of 5.86 *±* 1.77 years as reported in [4]. We used this follow-up time to assign the final vessel diameter 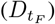 based on the growth rate and the initial diameter 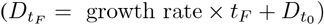.

This process yielded a cohort of *N* = 48 synthetic patients that reproduces the population-level statistics of the original clinical study while providing the complete state vectors required for modeling. Each synthetic patient was represented by the following vector 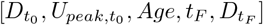. This sparse clinical data was used to fit the model parameters with the Bayesian strategy described below. Note that *Age* is not explicitly used in the model or during parameter estimation.

### C. Bayesian Inference Strategy

We framed the estimation of the unobservable damage parameters as a Bayesian inference problem. Let ***θ*** = {*k*_*dam*_, *k*_*e*_, *k*_*c*_, *K*_*σ*_, *k*_*D*_, *N*_*thresh*_} be the vector of unknown model parameters.

#### 1) Probabilistic Model

We assume the observed diameter *D*_*obs*_ at follow-up time *t*_*i*_ is normally distributed around the model prediction *D*_*model*_(*t*_*i*_, ***θ***):

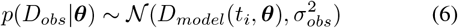

where *σ*_*obs*_ = 1 mm accounts for measurement uncertainty in clinical imaging.

#### 2) Priors

We use informative priors based on tissue engineering literature and physical constraints:

- **Rate Constants (***k*_*e*_, *k*_*c*_, *k*_*D*_): Log-uniform priors spanning orders of magnitude (e.g., 10^−11^ to 10^−7^ *s*^−1^) to assume no prior knowledge of the timescale.
- **Threshold (***N*_*thresh*_): Uniform prior 𝒰 [−4, 4] centered around the theoretical stability limit.
- **Damage Exponent (***k*_*dam*_): Uniform prior 𝒰 [1, 5] to capture the nonlinearity of the fatigue response.
- **Collagen Synthesis Gain (***K*_*σ*_): Uniform prior *𝒰* [0.05, 0.12] to capture the range used in previous work [15].

#### 3) Sampling and Validation

The posterior distribution *P* (***θ*** | *D*_*obs*_) was sampled using the Affine Invariant Markov Chain Monte Carlo (MCMC) Ensemble sampler (`emcee`) [22]. The inference engine was validated using a Leave-One-Out Cross-Validation (LOOCV) strategy, where the model was trained on *N* − 1 patients with Bayesian optimization using Gaussian processes [23] and tested on the held-out trajectory to assess predictive generalizability.

We compared the performance of our physics-based model in LOOCV against a simple linear regression model which used the initial patient parameters to predict the final vessel diameter at follow-up. Since *U*_*peak*,0_ was designed to be nosily correlated with the growth rate (Section II-B1), the linear regression model served as a necessary and informative baseline to confirm whether our physics-based approach performed, at least, comparably well.

### D. Representative trajectories and sensitivity

We use the fitted parameters to visualize representative trajectories of the stable, borderline, and unstable patient. We instantiate the following initial state vector at *t* = 0 for 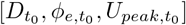 for each patient-type and solve for 7 years: 1) Stable: [28 mm, 0.35, 1.1 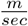 ], 2) Borderline: [36 mm, 0.30, 1.25 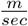], and 3) Unstable: [45 mm, 0.2, 1.35 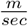].

We perform a sensitivity analysis around the Borderline patient by varying the values on the initial state vector by ± 30% to understand the behavior of the model near the bifurcation. We solve the system for 10 years and investigate diameter growth rates 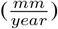 along with net % changes in elastin and collagen.

To inspect how changes in initial peak systolic velocity 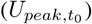 effect aneurysm progression, we inspect the trajectories of a set of Unstable patients of 3 sub-types who all have the same initial vessel diameter and elastin fraction 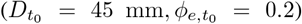 but three different 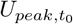 values corresponding to gradations of hypertension and elevated contractility: 1) Unstable–Type 1: 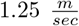, 2) Unstable–Type 2: 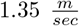, and 3) Unstable–T 3: 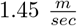.

Finally, to understand the effect of uncertainty from the Bayesian parameter estimation, we performed a sensitivity analysis for the Borderline patient using the confidence intervals for the estimates of the model parameters (Table I). We pseudo-randomly sampled 1,000 parameter combinations and ran a forward simulation for 7 years.

**TABLE I.** Posterior parameter estimates (median and 95% credible intervals) obtained from Bayesian inference. Parameter identifiability is quantified by the posterior contraction *S*, where *S* → 1 indicates strong data constraint.

| Parameter | Median | 2.5%ile | 97.5%ile | Identifiability ($S$) |
| --- | --- | --- | --- | --- |
| $k_e$ ($s^{-1}$) | $1.96 \times 10^{-8}$ | $8.51 \times 10^{-9}$ | $9.05 \times 10^{-8}$ | 0.92 (Strong) |
| $k_c$ ($s^{-1}$) | $5.14 \times 10^{-8}$ | $2.25 \times 10^{-8}$ | $9.54 \times 10^{-8}$ | 0.65 (Moderate) |
| $k_D$ ($m Pa^{-1} s^{-1}$) | $9.98 \times 10^{-10}$ | $2.32 \times 10^{-10}$ | $5.51 \times 10^{-9}$ | 0.48 (Moderate) |
| $N_{thresh}$ (-) | 0.92 | 0.12 | 1.56 | 0.81 (Strong) |
| $k_{dam}$ (-) | 2.40 | 1.24 | 3.22 | 0.55 (Moderate) |
| $K_\sigma$ (-) | 0.072 | 0.061 | 0.093 | 0.88 (Strong) |

## III. Results

### Parameter Recovery

Table I describes the posterior distributions for the model parameters after inference with the synthetic dataset. Parameter identifiability was quantified using the posterior contraction metric 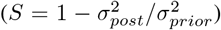. The parameters showed moderate-to-strong identifiability. We also test the sensitivity of the dynamics to parameter uncertainty in Figure 4 to confirm that behavior does not dramatically change within the uncertainty bounds.

### Leave-One-Out Validation

For LOOCV on the synthetic cohort, the physics-based model with parameters inferred through Bayesian optimization had a lower Root Mean Square Error (RMSE) for diameter prediction by ∼ 50% compared to the linear model (RMSE: 7.18 mm vs 14.51 mm, Pearson’s *r*: 0.849 vs 0.655). The physics-based model had more parameters, so performance cannot be directly compared. However, the physics-based model provides insight into internal dynamics and disease progression.

#### Trajectory Prediction and Bifurcation

The core contribution of this model is the prediction of non-linear trajectories and the details of the evolution of the disease. Figure 1 compares representative examples of Stable, Borderline, and Unstable patients. The model captures the accelerated growth observed in the Unstable patient, triggered when collagen remodeling is not adequately matching elastin loss (Figure 1A, B, and C, red line). The Unstable patient (red) undergoes some collagen deposition initially (C, years 0–4) which maintains pulse wave velocity (F) and increases wall stiffness (H), but damage accumulates through rapid elastin loss (B) with hemodynamic insult out-competing biological repair in later years (years 5–7). On the other hand, the Borderline patient (brown) undergoes significant collagen remodeling that increases pulse wave velocity and wall stiffness, maintaining elastin loss at levels comparable to the Stable patient. The Stable patient (blue) has little diameter growth along with age-related elastin loss and collagen remodeling.

**Fig. 1.**
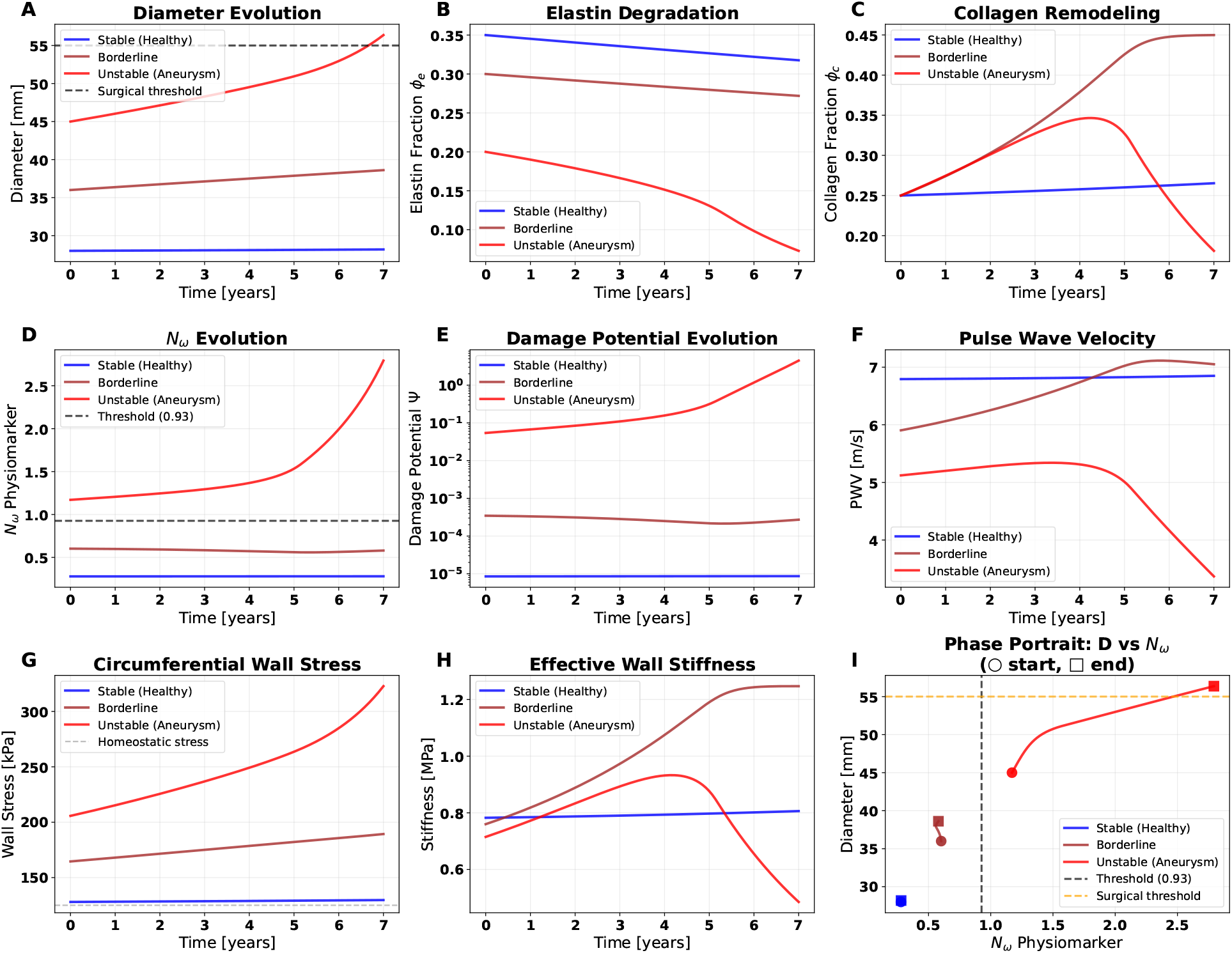
Predicted dynamics over 7 years for a Stable (blue), Borderline (brown), and an Unstable (red) patient using model parameters inferred through Bayesian estimation. The model distinguishes between the Borderline disease state that stabilizes via collagen remodeling (brown, A and C) from the Unstable disease state that suffers flutter-induced runaway expansion (red, A), accelerating elastin degradation (B), and collagen loss (C).

Figure 2 visualizes sensitivity analysis to initial conditions around the Borderline patient (Section II-D) by looking at 10-year outcomes in growth rate, percent change in elastin fraction and collagen fraction. Changes in initial elastin fraction and initial peak systolic velocity (A-C) with fixed initial vessel diameter had little effect on 10-year outcomes. Changing initial vessel diameter and initial peak systolic velocity (D-F) show clear regions of instability and bifurcation boundaries at the top-right corners. Changing initial elastin fraction and initial vessel diameter (G-I) also show changes in 10-year outcomes for high diameter initial conditions (*>* 45 mm). Figure 2F best shows the 3 regimes: 1) the top-right corner is the Unstable regime, 2) the dark-blue middle is Borderline, and 3) the light-blue bottom-band is Stable.

**Fig. 2.**
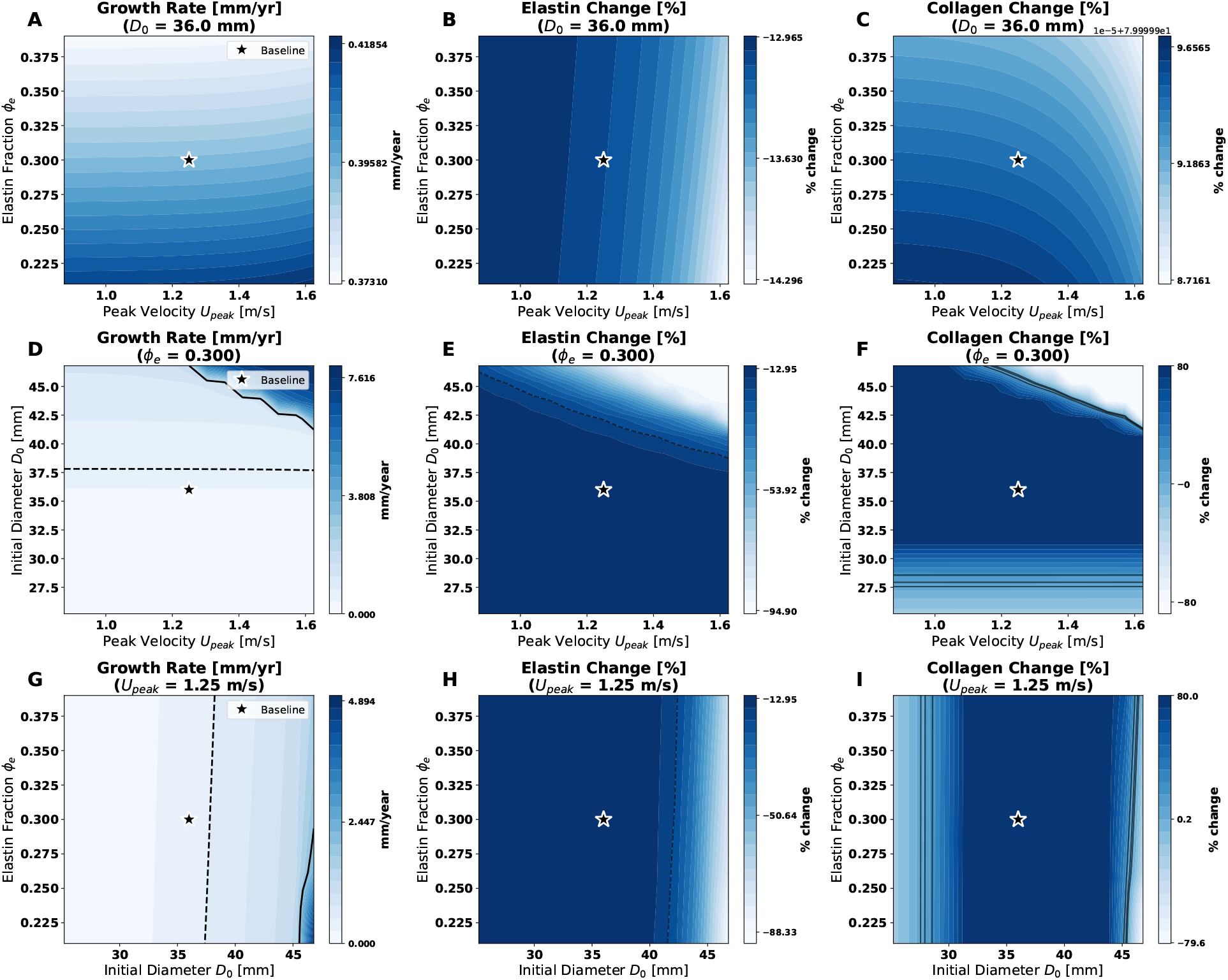
Sensitivity analysis around the Borderline patient from Figure 1 by varying initial parameters for vessel diameter, elastic fraction, and peak systolic blood velocity, [*D*_0_, *ϕ*_*e*,0_, *U*_*peak*,0_], by ± 30%. A-C) Initial vessel diameter is held constant at *D*_0_ = 36 mm. D-F) Initial elastin fraction is held constant at *ϕ*_*e*,0_ = 0.3. G-I) Peak systolic blood velocity is held constant at 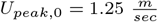 . The first column (A, D, G) shows vessel growth rate over 10 years. The second column (B, E, H) shows net % change in elastin fraction over 10 years. The last column (C, F, I) show net % change in collagen over 10 years. Note that the *color bar scales are separate* for each plot. The 3 regimes are clearly visible in panel F where the top-right corner is the Unstable regime, the dark-blue middle band is the Borderline regime, and the light-blue bottom band is the Stable regime.

Figure 3 shows the trajectories of the three Unstable subtypes with varying initial peak systolic velocities (*U*_*peak*,0_). All sub-types cross the surgical threshold within 10 years, but do so differently. Type 1 and 2 patients show initial increases in collagen fraction in response to stress. This transient collagen deposition temporarily limits vessel growth but ultimately proves insufficient to prevent aneurysm progression. In contrast, the Type 3 patient—with the highest *U*_*peak*,0_—has no compensatory collagen increase and proceeds rapidly through elastin and collagen loss, crossing the surgical threshold much earlier. This gradient of outcomes within the Unstable regime, shows how *U*_*peak*,0_ effects disease progression.

**Fig. 3.**
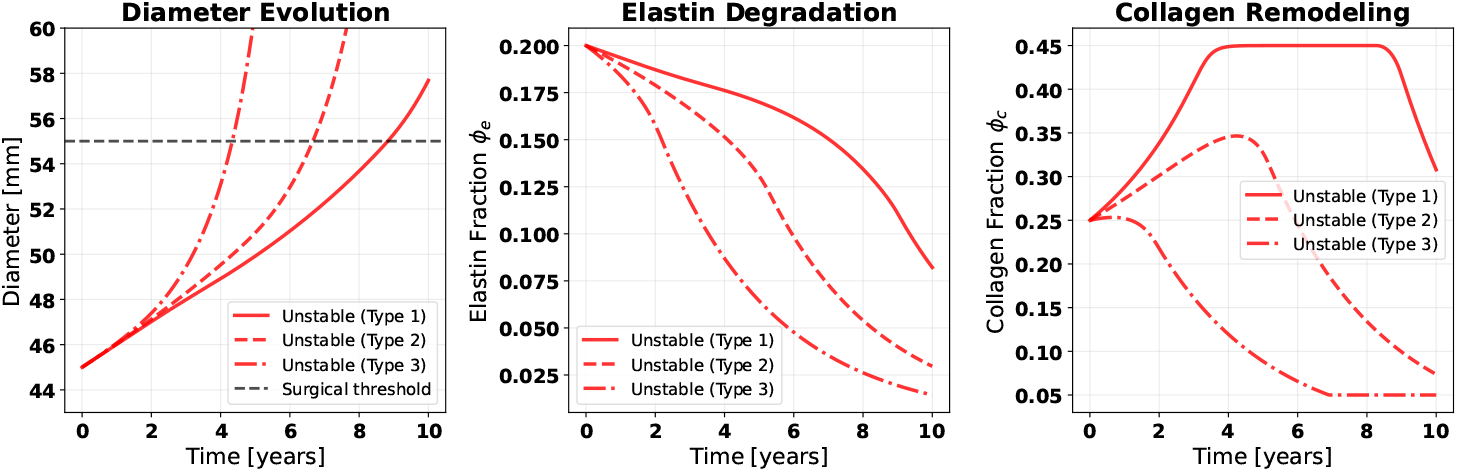
Predicted dynamics of 3 sub-types of Unstable patients with increasing initial peak systolic velocity (*U*_*peak*,0_) with Type 1 having the lowest value and Type 3 having the highest (Section II-D). All sub-types have dramatic elastin loss and cross the surgical threshold within 10 years. Type 1 and 2 have initial increases in collagen fraction but still suffer aneurysm later. Type 3 is at high risk and quickly proceeds with elastin and collagen loss, crossing the surgical threshold much earlier.

Figure 4 presents the sensitivity analysis for the Borderline patient using parameter uncertainty from Bayesian estimation. Despite sampling 1,000 parameter combinations from within the 95% confidence intervals, the model trajectories remained tightly bounded. Diameter evolution showed modest spread within the 95% confidence interval spanning approximately 5 mm at year 7. Elastin degradation and collagen remodeling trajectories were also robust to parameter uncertainty, suggesting that the inferred parameters provide reliable prognostic predictions.

**Fig. 4.**
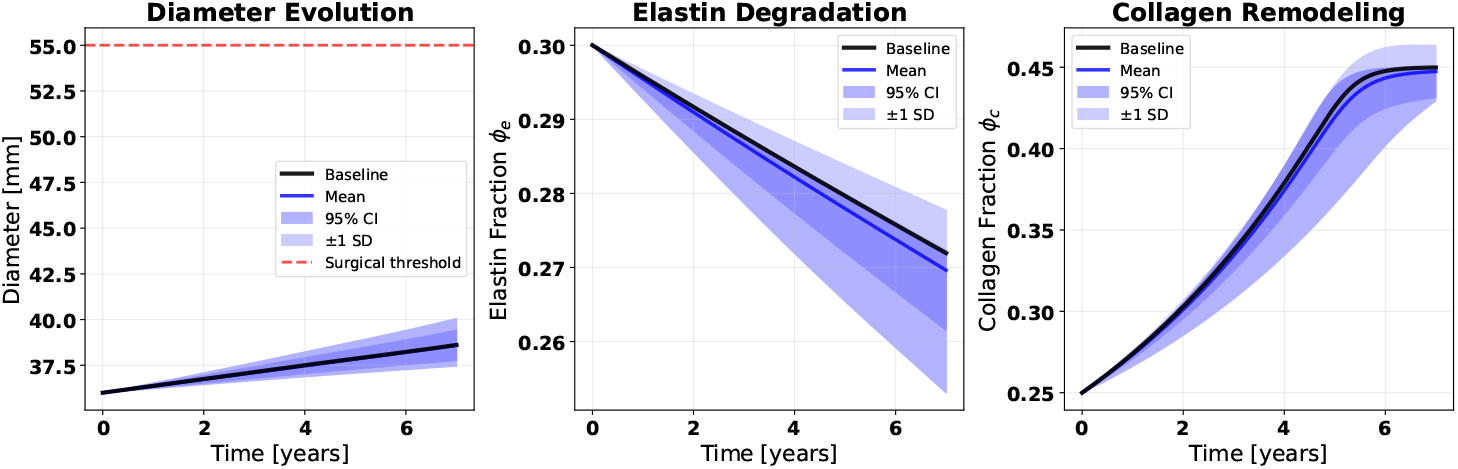
Sensitivity analysis performed with the confidence intervals of the Bayesian parameter estimates for the physics-based model around the Borderline patient through psuedo-random selection of 1,000 samples of parameter within 95% confidence intervals (Table I). Diameter evolution, elastin degradation, and collagen remodeling are not dramatically changed by the uncertainty in the parameter estimates.

## IV. Discussion

This work presents a proof-of-concept framework for linking the *N*_*ω*_ flutter physiomarker to tissue-level degradation dynamics. Using a Bayesian approach, we account for the inherent uncertainty in biological parameters while extracting prognostic information from sparse clinical measurements. While our results are derived from synthetic data and require validation on real patients, they demonstrate several properties that suggest the framework merits further development.

The “Damage Potential” formulation addresses a key limitation of linear stability analysis by translating the eigenvalue gap into a physical driving force for remodeling. The resulting bifurcation behavior aligns with clinical observations that aneurysms can remain stable for years before undergoing rapid expansion. This phenomenon is particularly evident in our Type 1 and Type 2 Unstable patients (Figure 3), who exhibit initial increases in collagen fraction that temporarily limit vessel growth, yet ultimately suffer progressive aneurysm development. The model captures this clinically observed latency period—the “calm before the storm”—that makes TAA particularly dangerous and difficult to manage with current surveillance methods.

The distinction between Borderline and Unstable trajectories offers important insights for clinical intervention. While initial vessel diameter is a non-modifiable risk factor, initial peak systolic velocity may be amenable to intervention through blood pressure management. Our sensitivity analysis (Figure 2D–F) demonstrates that reducing peak systolic velocity can shift patients from the Unstable regime into lowerrisk categories. For Borderline patients and for Types 1 and 2 Unstable patients, aggressive hypertension management could delay aneurysm progression or prevent it altogether.

This framework enables several clinically relevant applications. First, it provides a principled approach for predicting time-to-surgery, allowing clinicians to personalize surveillance intervals rather than relying on fixed imaging schedules. Patients predicted to remain stable could undergo less frequent imaging, while those on unstable trajectories could receive intensified monitoring. Second, the model could guide the intensity of hypertension management by quantifying how much reduction in peak systolic velocity would be required to shift a patient’s trajectory. Third, this framework could serve as a tool for evaluating treatment efficacy. By comparing pretreatment and post-treatment trajectories—for example, before and after 12 months of beta-blocker or doxycycline therapy— clinicians could objectively assess whether an intervention is producing the desired outcomes rather than waiting years to observe changes in diameter only. These applications are speculative until the framework is validated on real patient data.

### Future Work

The immediate priority is validation on real clinical data. An ideal validation cohort would include patients with serial 4D flow MRI or echocardiography (providing *U*_*peak*_ and *D* at multiple time-points) and, where available, tissue samples from surgical patients with measures of elastin and collagen fraction. Additionally, the damage model could be refined by incorporating results from in-vitro fatigue testing, and the spatial heterogeneity of wall properties could be addressed by extending to multi-compartment methods.

#### Limitations

We emphasize that this work should be viewed as a proof-of-concept establishing mathematical feasibility rather than a clinically validated tool. The most significant limitation is our reliance on synthetic data. While the cohort was calibrated to reproduce population-level statistics from clinical studies [4], synthetic patients cannot capture the full heterogeneity of real patient populations, including comorbidities, genetic factors, and measurement variability.

The damage potential formulation (Equation 2) is phenomenological. We justified the Softplus form based on mathematical convenience and physical intuition, but the true relationship likely involves complex mechanotransduction pathways. Future work could incorporate mechanistic models informed by fatigue testing.

Parameter identifiability remains a concern. While some parameters showed strong posterior contraction (*S >* 0.8), others had only moderate identifiability (*S* ∼ 0.5), suggesting that additional measurements may be needed. Moreover, because initial peak systolic velocities were assigned to correlate with growth rates, the improvement over linear regression, while encouraging, requires further confirmation. Finally, the model assumes elastin is non-regenerative in adults, which is the consensus view but may not hold universally.

## V. Conclusion

We developed a proof-of-concept system identification framework for aortic aneurysm progression. By coupling *N*_*ω*_ to elastin degradation and collagen remodeling, we establish the mathematical foundation for making a static diagnostic criterion into a dynamic prognostic tool. The Bayesian inference approach enables estimation of parameters from sparse data while quantifying uncertainty. Using synthetic patients calibrated to clinical statistics, we demonstrated that this physics-informed framework can distinguish between self-stabilizing and runaway disease trajectories and capture the clinically observed latency period before rapid expansion. While validation on real patient data remains essential, this work suggests that physics-based modeling of flutter-induced damage offers a promising pathway toward personalized risk stratification.

## Data Availability

All data produced in the present study are available upon reasonable request to the authors

## Notes

### Competing Interest Statement

The authors have declared no competing interest.

### Funding Statement

This study did not receive any funding

